# Estimated effects of plant- and animal-based protein food strategies on muscle function, strength and lean mass after 6 years: a target trial emulation in the Canadian Longitudinal Study on Aging

**DOI:** 10.64898/2026.11.20.26368611

**Authors:** Didier Brassard, Andrea J Glenn, Stéphanie Chevalier, Matthieu Ruiz, Jun Li, Anne-Julie Tessier

**Affiliations:** Montreal Heart Institute, and Department of Nutrition, Université de Montréal, Montréal, QC H1T 1C8, Canada; Department of Medical Biology, Université du Québec à Trois-Rivières, Trois-Rivières, G8Z 4M3, Canada; Department of Nutrition and Food Studies, New York University, New York, NY 10003, United States; Department of Nutrition, Harvard T.H. Chan School of Public Health, Boston, MA 02115, United States; School of Human Nutrition, McGill University, Montreal, H3A 0G4, Canada; Research Institute of the McGill University Health Centre, Montreal, H4A 0B1; Brigham and Women’s Hospital, Boston, MA, 02115, United States

**Keywords:** older adults, muscle, CLSA, causal inference, target trial emulation, nutritional epidemiology, plant protein foods, animal protein foods

## Abstract

**Background:** Dietary guidelines emphasize the consumption of plant-based protein foods for cardiovascular health. However, it remains unclear how the balance of plant- and animal-based protein food consumption contributes to muscle health.

**Objective:** Our objective was to estimate the effect of increasing the consumption of plant- (PLANT) relative to animal-based (ANIMAL) protein foods on physical function, including muscle function, strength, and lean mass, in middle-aged and older adults. We also examined the effects of increasing both protein sources (BOTH), within a healthy dietary pattern (HEALTHY), or limiting animal protein foods consumption (HEALTHY-P+).

**Methods:** Non-experimental data from the Canadian Longitudinal Study on Aging (CLSA) comprehensive cohort were used to emulate a target trial. Middle-aged and older adults were followed for 6 years. Sociodemographic and diet data (frequency via the Short Diet Questionnaire) were assessed every three years. Objectively measured outcomes were muscle function (Timed Up&Go, 4-m walking speed, chair rise time), appendicular lean soft tissue (ALST) by dual-energy X-ray absorptiometry and handgrip strength. The parametric g-formula was used to account for time-varying confounding and loss to follow-up.

**Results:** A total of 21,636 participants were eligible (median [Q1-Q3] age, 62 [54-70] years; 49% female). Across all hypothetical interventions, the HEALTHY intervention consistently improved outcomes. Compared with no intervention, HEALTHY yielded the largest estimated differences: Timed Up&Go (−0.15 s; 95%CI: −0.24, −0.07), chair rise time (−0.03 s; 95%CI: −0.06, −0.01), handgrip strength (+0.5 kg; 95%CI: 0.3, 0.7), and ALST (+0.12 kg, 95%CI: 0.05-0.18). BOTH showed consistent but smaller effect estimates. Further limiting animal protein did not confer additional benefits. Interventions increasing PLANT or ANIMAL alone showed 95%CIs compatible with no effect. Effect estimate magnitudes were below clinically important differences.

**Conclusion:** In middle-aged and older adults, findings suggest that a balanced approach to increasing consumption of both animal and plant-based protein foods may yield modest benefits for muscle health compared with no intervention, particularly within a healthy dietary pattern.

## 1. Introduction

Dietary guidelines emphasize plant-based foods such as nuts, seeds, legumes, unsaturated fats, vegetables and fruits and whole grains for cardiovascular health (1–3). For example, Canada’s Food Guide recommends consuming more plant-based protein foods than animal-based protein foods as well as a diet rich in vegetables and fruits and whole grains (1). In observational studies, adhering to Canada’s Food Guide recommendations reduced the 10-year cardiovascular disease risk by 24% and improved heart age (4, 5). Similarly, a meta-analysis of 24 cohorts support consuming more plant-based than animal-based foods for cardiovascular disease prevention (6).

Yet, healthy aging encompasses more than chronic disease and cardiovascular risk prevention. The World Health Organization provides a definition of healthy aging, which is the process of developing and maintaining the functional ability that enables wellbeing in older age (7–9). A direct contributor to functional ability is intrinsic capacity, defined as the composite of all the physical and mental capacities that an individual can draw on (8). In other words, the ability to maintain physical function as well as mobility are key determinants of healthy aging (7). Observational studies showed associations between dietary patterns rich in vegetables, fruits, whole grains, unsaturated fats, nuts, legumes, as well as low-fat dairy products, and healthy aging (10). However, it is unclear how the relative proportion of plant- and animal-based protein foods contribute to physical function among middle-aged and older adults. While emphasizing plant-based protein foods relative to animal-based protein foods has demonstrated benefits for chronic disease prevention (4–6, 10, 11), animal-based protein foods may also contribute to nutrient intake adequacy, optimal protein intakes and greater muscle health among aging populations (12–14).

The long-term contributions of animal- and plant-based protein foods to optimal muscle function, strength and mass have not been well characterized in Canadian populations. Furthermore, it is unclear how strategies that limit animal-based protein foods affect muscle health or whether consuming these protein foods as part of a broader healthy dietary pattern, such as vegetables and fruits or whole grains, changes their effects. Thus, our objective was to estimate differences in muscle function and strength, and lean mass under hypothetical dietary interventions that modify the frequency of consumption of plant- and animal-based protein foods, alone or within a healthy dietary pattern, among middle-aged and older adults. More precisely, compared with no intervention, we examined the effect of six intervention strategies that increase the frequency of consumption of: 1) plant-based protein foods; 2) animal-based protein foods; 3) both plant- and animal-based protein foods; 4) only plant-based protein foods, while limiting animal-based protein foods; 5) both plant- and animal-based protein foods, but also vegetables and fruits and whole grains; 6) same as 5, but limiting animal-based protein foods.

## 2. Methods

### 2.1 Study Design and Participants

Data from the Canadian Longitudinal Study on Aging (CLSA) comprehensive cohort were used for the present study. Complete details about the CLSA have been reported (15, 16). A total of 30,097 community-dwelling participants aged 45-85 years were recruited between 2010 and 2015 among seven provinces in Canada. Participants were randomly selected but had to live within 50 km of 11 data collection sites, had to speak French or English, and had to be free of cognitive impairment that would have precluded providing informed consent (16). For the present study, participants attended two follow-up visits after three and six years. At each visit, participants underwent comprehensive examination, including sociodemographic, lifestyle and health questionnaires, physical examination, body composition, blood sampling, as well as muscle strength and function assessments. Participants with complete covariate and dietary assessment data at baseline were eligible. They were excluded if they had multiple sclerosis, Alzheimer’s or Parkinson’s disease, had a surgery in the last three months, had polio, had chemotherapy in the last four weeks, had a traumatic brain injury with memory problems, had post-traumatic stress disorder or dialysis at baseline. The exclusion criteria were selected considering their potential impact on diet as well as muscle strength and functions outcomes.

Given the causal inference nature of the research objective, the analysis aimed to emulate a target trial (17). **Supplemental Table 1** presents the target trial emulation specification and underlying assumptions. The use of data from the CLSA for this project was approved by the Montreal Heart Institute ethics committee (#2025-3517).

### 2.2 Outcome

**Supplemental Table 2** describes the source of data and details of assessment methods for each outcome. The outcomes were Timed Up&Go, average time for one chair rise, walking speed over a 4-metre course, handgrip strength, and appendicular lean soft tissue (ALST) measured using dual-energy X-ray absorptiometry (DXA; Hologic Discovery A). Handgrip strength was measured as the highest value of 3 trials using a handheld dynamometer (Tracker Freedom Wireless Grip). ALST was calculated as the sum of the upper and lower limbs lean mass minus bone mineral content and considered as a proxy for appendicular skeletal muscle mass. Outcomes were measured using standardized procedures across the 11 data collection sites (18).

### 2.3 Exposure

#### 2.3.1 Diet frequency

Diet in the previous 12 months was assessed using the interviewer-administered Short Diet Questionnaire (SDQ) (19, 20). The SDQ was developed based on the Canadian self-administered 78-item semi-quantitative food-frequency questionnaire (21). At baseline, the SDQ assessed the frequency of consumption of 30 foods and 6 beverages, without portion size estimation. Based on a subsample of 232 self-selected participants from the CLSA, the test-retest reliability of the SDQ was evaluated (19). Intraclass correlation coefficients for item combination were fair (range, 0.35 to 0.72). The highest intraclass correlation coefficients were for yoghurt (0.72), eggs (0.70), and milk (0.62), while the lowest were for total meat with processed meat (0.35), cheese (0.42) and calcium supplemented foods (0.44). Information on dietary supplements (vitamins, minerals) or natural health products use were collected as part of the other questionnaire during in-home interviews. After baseline, 5 additional questions were added to the SDQ to assess the frequency of consumption of packaged foods, fruit drinks, sugar-sweetened beverages, artificially sweetened beverages, and energy drinks.

In the present study, frequency variables were grouped into 7 categories similar to the Canada’s Food Guide food categories (1). The categories were vegetables and fruits, whole grain foods, plant-based protein foods (including plant-based beverages), animal-based protein foods (including dairy and milk), low nutritive value foods, fat sources, other beverages and sugary drinks. One variable assessing the consumption of calcium-fortified foods was unclassified. SDQ data with missing data for more than 5 variables were considered incomplete.

##### Hypothetical dietary intervention strategy

Hypothetical dietary intervention strategies were designed to estimate the effect of modifying the frequency of consumption of plant-based protein foods, animal-based protein foods, or both, and as part of a healthy eating strategy consistent with Canada’s Food Guide (CFG) recommendations (1). **Table 1** presents the daily target of the frequency of consumption of key food categories under each strategy. The targets were derived based on diet simulations generated by Health Canada (22). The simulations provide the minimal number of foods to consume to meet both nutrient intake adequacy and adhere to CFG recommendations (22). In the present study, the least restrictive standards (females aged 71 y or more) were used to derive frequency targets consistent with a healthy eating pattern. These standards were further revised to be capped at the 90^th^ percentile of consumption in Canada (23), yielding the target in Table 1. All strategies aimed to estimate the effect of consuming a minimum frequency except “*Higher plant but limited animal protein foods*” and “*Higher plant but limited animal protein foods within a healthy dietary pattern*”. The latter two strategies include a maximal frequency of 1.5 times per day for animal-based protein foods. In the two interventions including targets for vegetables and fruits and whole-grain foods (“with healthy diet”), the target frequency of consumption for plant-based protein foods and animal-based protein foods were set at 1.6 and 1.5 times per day, respectively. These frequencies reflect the CFG recommendation to consume plant-based protein foods more frequently than animal-based protein foods (1).

**Table 1:** Hypothetical intervention targets for the daily frequency of food consumption at all follow-ups among eligible participants CLSA participants (n=21,636)^1^.

| **Intervention** | **Causal question: compared with no intervention, what would have been the outcome had all participants …** | **Plant-based protein foods<sup>*2*</sup>** | **Animal-based protein foods<sup>*3*</sup>** | **Vegetables and fruits** | **Whole-grain foods** |
|---|---|---|---|---|---|
| No intervention | --- | No intervention |  |  |  |
| Higher plant protein foods | consumed **more plant-based** protein foods? | 1.5 or more | --- | --- | --- |
| Higher animal protein foods | consumed **more animal-based** protein foods? | --- | 2.5 or more | --- | --- |
| Higher total protein foods | consumed **more plant- and animal-based protein foods**? | 1.5 or more | 1.5 or more | --- | --- |
| Higher plant but limited animal protein foods | consumed **more plant- but limited animal-based protein foods**? | 1.5 or more | 1.5 or less | --- | --- |
| Higher total protein foods within a healthy dietary pattern | consumed **more plant- and animal-based protein foods, as part of a healthy diet**? | 1.6 or more | 1.5 or more | 3.5 or more | 1 or more |
| Higher plant but limited animal protein foods within a healthy dietary pattern | consumed **more plant-based protein foods, but limited animal-based protein foods, as part of a healthy diet**? | 1.6 or more | 1.5 or less | 3.5 or more | 1 or more |

<sup>*1*</sup>In the hypothetical intervention, participants would be instructed to consume recommended foods as frequently as specified in the table. For example, in the *Higher plant protein foods* intervention, a participant that would report consuming plant-based protein foods **less** frequently than 1.5 times per day would receive dietary counselling to **increase** their frequency of consumption to 1.5 times per day. To maintain energy balance, the participant would also be instructed to **decrease** their frequency of consumption of foods not recommended proportionately to the extent of the increase. Participants reporting consuming plant-based protein foods more frequently than 1.5 times per day would be instructed to maintain their consumption. Finally, to avoid consuming more foods, the participants would be instructed to decrease their intake of foods not recommended proportionately to the increase required to meet the intervention threshold of foods recommended. Foods not recommended include sugary beverages and low nutritive value foods.
<sup>*2*</sup>Plant-based beverages are included in plant-based protein foods.
<sup>*3*</sup>Milk and dairy foods are included in animal-based protein foods.

### 2.4 Covariates

Covariates were selected to address confounding at baseline, throughout follow-up, and reduce selection bias due to individual and sociodemographic factors, disease burden and propensity towards health-seeking behaviors. The baseline value of the outcome was included to account for the effect of dietary habits and lifestyle prior to baseline. We assumed that 1) the baseline value of outcome is a mediator of the effect of diet on end of follow-up value of outcome (24) and 2) there is no confounding between the baseline value of outcome (i.e., the assumed mediator) and the end of follow-up value of outcome (i.e., the outcome of interest), e.g., anabolic resistance or unknown “robustness” factors not completely explained by the other covariates. Covariates were age, sex, height, baseline value of outcome, education level, province of residence, supplement use (vitamin, mineral, omega 3-6-9, or natural health product), former smoking, frequency of added fats, total frequency of food consumption, body weight, menopausal status (female only), hormone replacement therapy use (female only), marital status, frequency of social participation, living alone, retirement status, high blood pressure, diabetes, cancer diagnosis, current smoker, frequency of alcohol consumption in the past 12 months, sitting time in the past week, as well as light intensity or greater physical activity in the past week. All covariates were collected using standardized interviewer-administered questionnaires and procedures (25). Physical activity was measured using the Physical Activity Scale for the Elderly (PASE) questionnaire (26, 27). Of note, the SDQ does not collect detailed information on portion size. Thus, to account for total food intake as covariate, the sum of all frequencies of food and beverage consumption was used as a proxy (“total frequency of food consumption”). The adjustment for height, body weight and physical activity was considered as an alternative to total energy intake adjustment (28).

### 2.5 Statistical Analyses

All analyses were conducted using R version 4.4.3 (2025-02-28) (R foundation) and the package gfoRmula v1.1.1.

#### Main analysis

The main analysis involved emulating a sustained intervention strategy where participants would be instructed to adhere to pre-specified dietary targets indicated in Table 1. The parametric g formula was used to address confounding, selection bias, and emulate the sustained intervention strategy (29). First, the values of all time-varying variables across baseline and follow-ups were predicted using parametric models, while adjusting for baseline covariates and the remaining time-varying variables. Second, copies of the data were generated using Monte Carlo simulations and predicted values. Each copy corresponded to a hypothetical intervention where the reported frequencies of food consumption were modified (hypothetically “intervened on”) to correspond to the prespecified targets. Third, the outcome model was fitted using predicted values from one prior follow-up, covariates, and diet variables. Then, the mean outcome value was estimated under each hypothetical intervention and compared to the value of outcome when dietary intakes are left unchanged, yielding the end of follow-up mean difference in outcome. Finally, variance was estimated using 500 bootstrap resamples and the percentile method.

Furthermore, to mitigate confounding by disease burden, participants were “excused” from adhering to the intervention at follow-up 1 and 2 when developing conditions listed as exclusion criteria. In other words, the diet of participants was left unchanged under the hypothetical intervention while their data were kept for the outcome model. Supplemental Table 3 shows the modelling approach for each variable.

#### Subgroup and Sensitivity Analyses

For subgroup analyses, data were stratified by sex and age (<65 y, 65 y or older) to assess potential differences. Sensitivity analyses were conducted to challenge key assumptions of causal inference estimation. First, the probability of injury at the end of follow-up was used as a negative control outcome (30). Injuries serious enough to limit normal activities in the past 12 months were considered (e.g., road accident, struck by an object, fall from a height, natural factors), but fall from same level were excluded. The probability of injury should not be strongly related to dietary habits in absence of unmeasured or residual confounding. Second, the values of time-varying variables predicted by models in the g formula algorithm were compared to observed values to verify model misspecification (31). Both predicted and observed values should be similar when data are modelled correctly.

## 3. Results

### 3.1 Participants

Of the 30,097 participants in the CLSA comprehensive cohort, 21,636 met the eligibility criteria (**Figure 1**). The median age of eligible participants was 62 years (Q1-Q3: 54-70) and 49% were female (**Table 2**). Characteristics of eligible and excluded CLSA participants (n=8461) are provided in **Supplemental Table 4**. Among eligible participants, 4,209 (19.5%) were lost during follow-up. Of the 17,427 participants not otherwise lost to follow-up, 7,243 (41.6%) could not attend the final examination because of COVID-19 restrictions.

**Figure 1:**
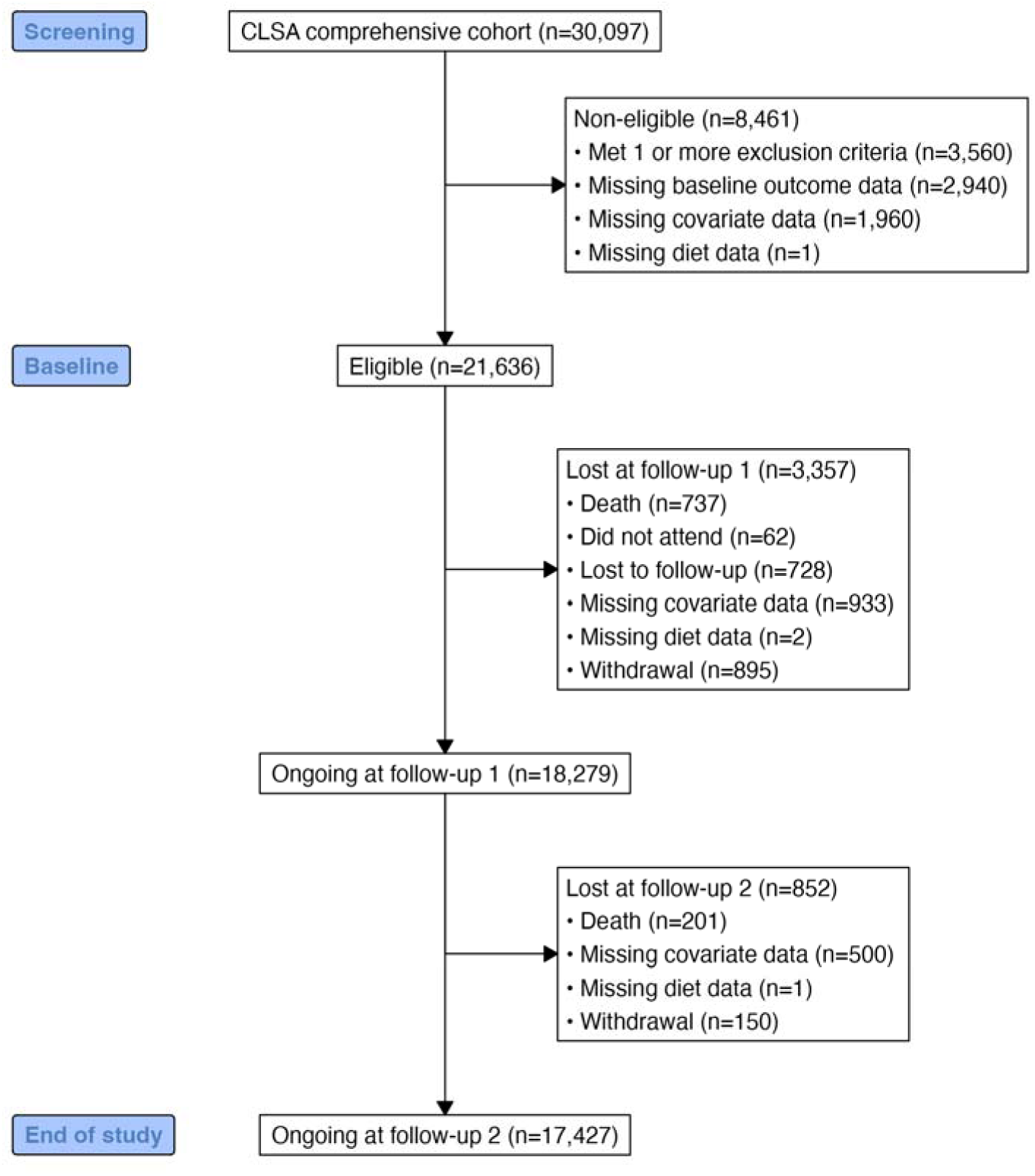
Flowchart of participants among CLSA participants.

**Table 2:**
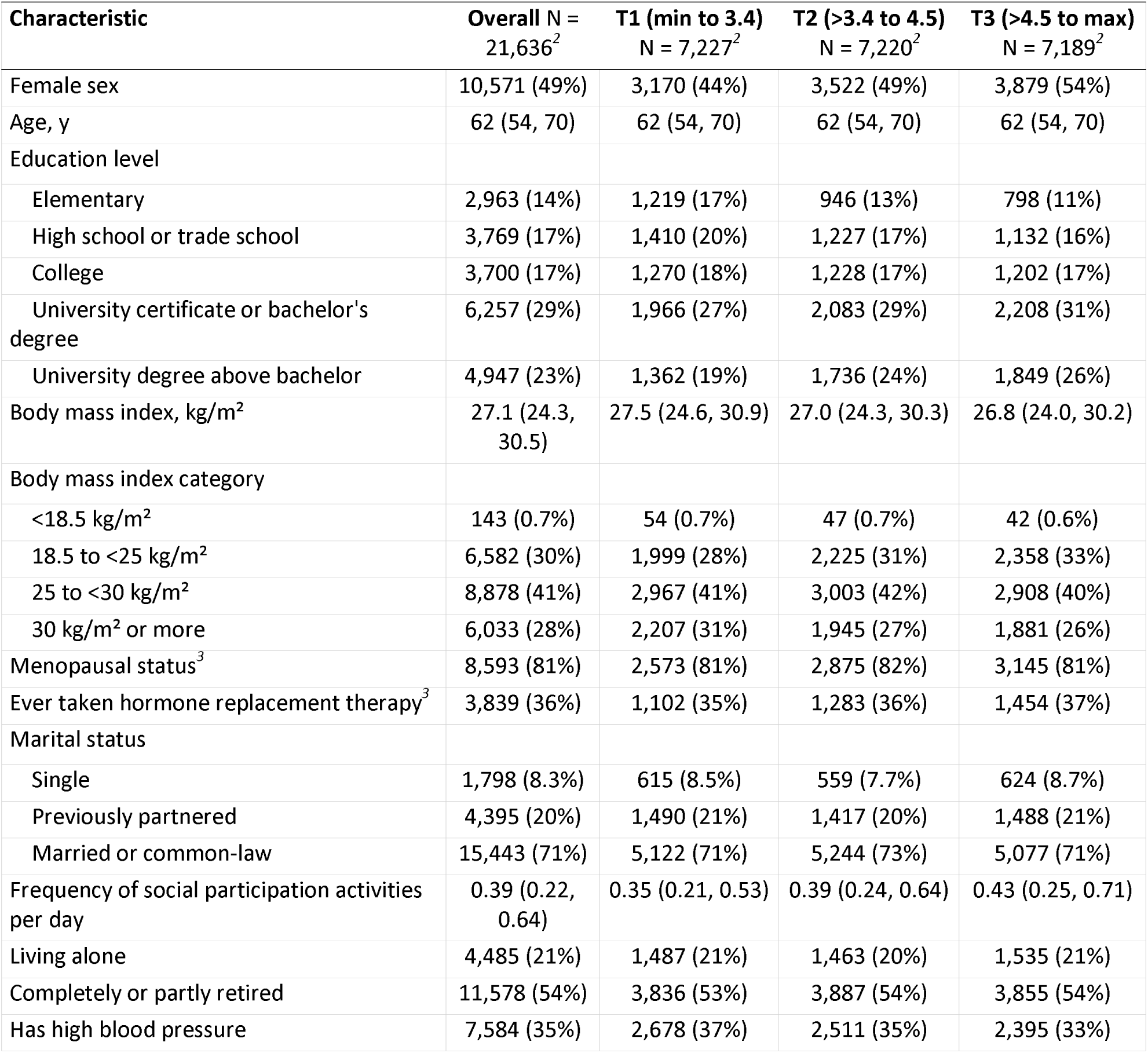
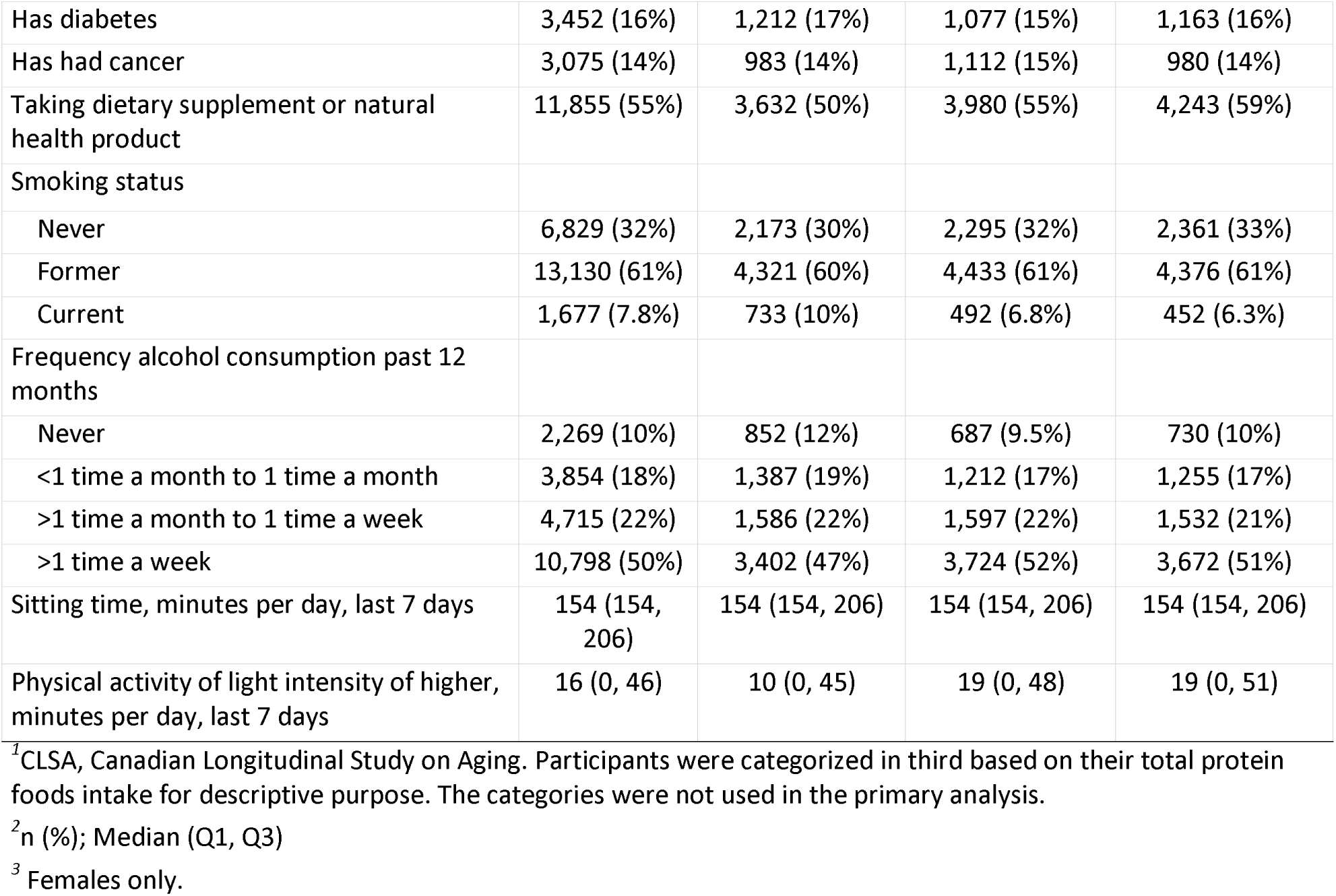
Characteristics of eligible CLSA participants at baseline across tertiles of protein foods frequency per day^1^.

| **Characteristic** | **Overall** N = 21,636<sup>2</sup> | **T1 (min to 3.4)** N = 7,227<sup>2</sup> | **T2 (>3.4 to 4.5)** N = 7,220<sup>2</sup> | **T3 (>4.5 to max)** N = 7,189<sup>2</sup> |
|---|---|---|---|---|
| Female sex | 10,571 (49%) | 3,170 (44%) | 3,522 (49%) | 3,879 (54%) |
| Age, y | 62 (54, 70) | 62 (54, 70) | 62 (54, 70) | 62 (54, 70) |
| Education level |  |  |  |  |
| Elementary | 2,963 (14%) | 1,219 (17%) | 946 (13%) | 798 (11%) |
| High school or trade school | 3,769 (17%) | 1,410 (20%) | 1,227 (17%) | 1,132 (16%) |
| College | 3,700 (17%) | 1,270 (18%) | 1,228 (17%) | 1,202 (17%) |
| University certificate or bachelor's degree | 6,257 (29%) | 1,966 (27%) | 2,083 (29%) | 2,208 (31%) |
| University degree above bachelor | 4,947 (23%) | 1,362 (19%) | 1,736 (24%) | 1,849 (26%) |
| Body mass index, kg/m² | 27.1 (24.3, 30.5) | 27.5 (24.6, 30.9) | 27.0 (24.3, 30.3) | 26.8 (24.0, 30.2) |
| Body mass index category |  |  |  |  |
| <18.5 kg/m² | 143 (0.7%) | 54 (0.7%) | 47 (0.7%) | 42 (0.6%) |
| 18.5 to <25 kg/m² | 6,582 (30%) | 1,999 (28%) | 2,225 (31%) | 2,358 (33%) |
| 25 to <30 kg/m² | 8,878 (41%) | 2,967 (41%) | 3,003 (42%) | 2,908 (40%) |
| 30 kg/m² or more | 6,033 (28%) | 2,207 (31%) | 1,945 (27%) | 1,881 (26%) |
| Menopausal status<sup>3</sup> | 8,593 (81%) | 2,573 (81%) | 2,875 (82%) | 3,145 (81%) |
| Ever taken hormone replacement therapy<sup>3</sup> | 3,839 (36%) | 1,102 (35%) | 1,283 (36%) | 1,454 (37%) |
| Marital status |  |  |  |  |
| Single | 1,798 (8.3%) | 615 (8.5%) | 559 (7.7%) | 624 (8.7%) |
| Previously partnered | 4,395 (20%) | 1,490 (21%) | 1,417 (20%) | 1,488 (21%) |
| Married or common-law | 15,443 (71%) | 5,122 (71%) | 5,244 (73%) | 5,077 (71%) |
| Frequency of social participation activities per day | 0.39 (0.22, 0.64) | 0.35 (0.21, 0.53) | 0.39 (0.24, 0.64) | 0.43 (0.25, 0.71) |
| Living alone | 4,485 (21%) | 1,487 (21%) | 1,463 (20%) | 1,535 (21%) |
| Completely or partly retired | 11,578 (54%) | 3,836 (53%) | 3,887 (54%) | 3,855 (54%) |
| Has high blood pressure | 7,584 (35%) | 2,678 (37%) | 2,511 (35%) | 2,395 (33%) |
| Has diabetes | 3,452 (16%) | 1,212 (17%) | 1,077 (15%) | 1,163 (16%) |
| Has had cancer | 3,075 (14%) | 983 (14%) | 1,112 (15%) | 980 (14%) |
| Taking dietary supplement or natural health product | 11,855 (55%) | 3,632 (50%) | 3,980 (55%) | 4,243 (59%) |
| Smoking status |  |  |  |  |
| Never | 6,829 (32%) | 2,173 (30%) | 2,295 (32%) | 2,361 (33%) |
| Former | 13,130 (61%) | 4,321 (60%) | 4,433 (61%) | 4,376 (61%) |
| Current | 1,677 (7.8%) | 733 (10%) | 492 (6.8%) | 452 (6.3%) |
| Frequency alcohol consumption past 12 months |  |  |  |  |
| Never | 2,269 (10%) | 852 (12%) | 687 (9.5%) | 730 (10%) |
| <1 time a month to 1 time a month | 3,854 (18%) | 1,387 (19%) | 1,212 (17%) | 1,255 (17%) |
| >1 time a month to 1 time a week | 4,715 (22%) | 1,586 (22%) | 1,597 (22%) | 1,532 (21%) |
| >1 time a week | 10,798 (50%) | 3,402 (47%) | 3,724 (52%) | 3,672 (51%) |
| Sitting time, minutes per day, last 7 days | 154 (154, 206) | 154 (154, 206) | 154 (154, 206) | 154 (154, 206) |
| Physical activity of light intensity of higher, minutes per day, last 7 days | 16 (0, 46) | 10 (0, 45) | 19 (0, 48) | 19 (0, 51) |

<sup>*1*</sup>CLSA, Canadian Longitudinal Study on Aging. Participants were categorized in third based on their total protein foods intake for descriptive purpose. The categories were not used in the primary analysis.
<sup>*2*</sup>n (%); Median (Q1, Q3)
<sup>*3*</sup> Females only.

Compared with participants of the lowest third of total protein food intake, participants in the highest third included a greater proportion of females, participants with a university education, participants reporting taking dietary supplement or natural health product and a lower proportion of current smokers. The median (Q1-Q3) follow-up time was 5.7 years (5.3-6.0). **Supplemental Table 5** shows the number of participants with missing diet assessment and covariates across follow-ups. The proportion of participants meeting the intervention thresholds and the mean outcome values observed across follow-ups are reported in **Supplemental Tables 6 and 7**, respectively.

### 3.2 Main Analysis

Among the hypothetical interventions, consuming higher total protein foods (plant- and animal protein food targets: ≥1.6 and ≥1.5 times/day, respectively) within a healthy dietary pattern intervention would have resulted in improvement in health outcomes across all five measures (**Figure 2**). After 6 years, the estimated mean differences comparing this intervention with no intervention were −0.15 s for Timed Up&Go (95%CI: −0.24, −0.07), −0.03 s for one chair rise (95%CI: −0.06, −0.01), +0.5 kg for handgrip strength (95%CI: 0.3, 0.7). The intervention of increasing higher total protein foods alone would have also yielded favorable outcomes across all measures, except for walking speed. Compared with no intervention, the estimated mean differences were − 0.06 s for Timed Up&Go (95% CI: −0.13, 0.00) and −0.02 s for one chair rise (95% CI: − 0.05, 0.00). For ALST, the 95%CI of estimated end-of-follow-up mean differences ranged from −0.10 to 0.18 kg across interventions. The largest estimated difference was for the intervention of consuming higher total protein foods within a healthy dietary pattern (+0.12; 95%CI: 0.05 to 0.18; Figure 2).

**Figure 2:**
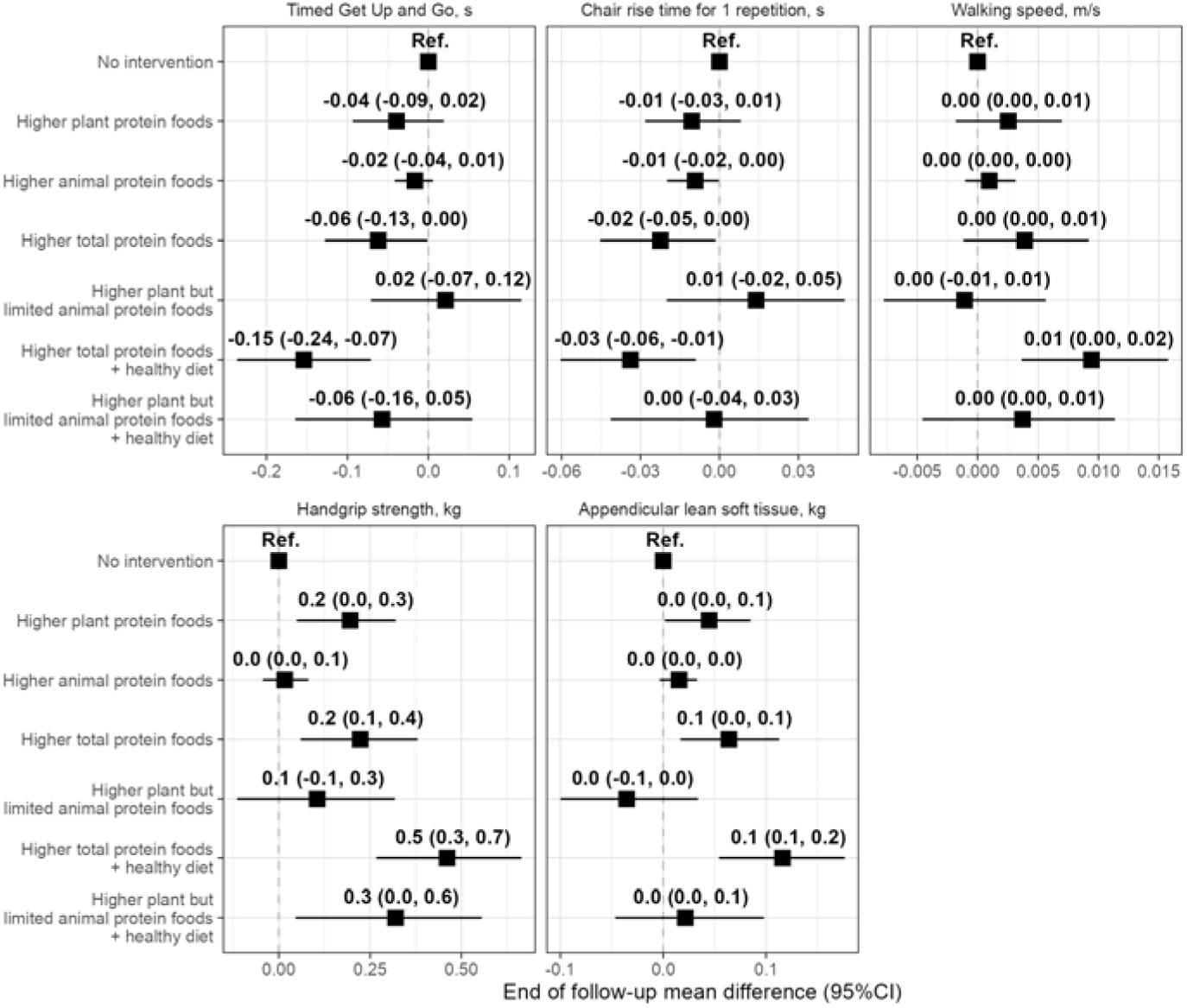
End-of-follow-up mean differences (95% CI) in muscle function, handgrip strength and appendicular lean soft tissue under each 6-year hypothetical dietary interventions compared with no intervention among eligible CLSA participants (n=21,636). Effect estimates were obtained using the parametric g-formula algorithm. Models were adjusted for the covariates age, sex, height, baseline value of outcome, education level, province of residence, supplement use (vitamin, mineral, omega 3-6-9, or natural health product), former smoker, protein foods, plant-based, protein foods, animal-based, fat sources, total foods, bodyweight, menopausal status (female only), hormone replacement therapy use (female only), marital status, social participation, living alone, retirement status, high blood pressure, diabetes, cancer diagnosis, current smoker, frequency of alcohol consumption in the past 12 months, sitting time in the past week, as well as light intensity or greater physical activity in the past week. CLSA, Canadian Longitudinal Study on Aging

Compared with no intervention, the intervention strategy that increased plant-based protein foods alone would have resulted in 0.2 kg increase in handgrip strength at the end of follow-up (95% CI: 0.0, 0.3), whereas differences in physical function and ALST were close to the null. The intervention that increased animal-based protein foods alone would have resulted in little or no difference across outcomes. Similarly, consuming higher plant protein foods while limiting animal protein foods would have resulted in no difference across most outcomes (Figure 2). The only exception was a greater handgrip strength (+0.3 kg; 95%CI, 0.0, 0.6) when this strategy was to be combined within a healthy dietary pattern.

### 3.3 Subgroup Analysis

**Supplemental Figures 1 and 2** show results stratified by sex and age (<65, 65 y or older), respectively. The estimated effects were consistent in direction and size in males and females, except for handgrip strength. At the end of follow-up, the estimated mean differences in handgrip strength were all slightly greater among males than among females compared with no intervention. For example, the intervention that increased total protein foods within a healthy dietary pattern would have improved handgrip strength by 0.8 kg (95%CI: 0.5, 1.1) in males, but only 0.1 kg (95%CI: −0.1, 0.4) in females, resulting in a 0.6 kg (95%CI: 0.3, 1.1) greater increase. For the same intervention, the estimated difference in Timed Up&Go was compatible with a 0.11 second faster time (95%CI: −0.29, 0.07) in participants aged 65 y or older compared with participants aged less than 65 y.

### 3.4 Sensitivity Analysis

The estimated probability of serious injury during the past 12 months did not reveal strong bias. The 95%CI were compatible with both a small decrease to a small increase of the injury probability, including the null value (e.g., relative probability, 0.95; 95%CI: 0.88, 1.03 for higher total protein foods within a healthy dietary pattern compared with no intervention; **Supplemental Table 8**). However, in the sex-stratified analysis, the intervention that increased animal protein foods only was associated with an increased probability of injury in females (relative probability, 1.04; 95%CI: 1.01, 1.08). For model misspecification, the g-formula predicted values for time-varying diet and covariates were generally consistent with observed values (**Supplemental Figure 3**).

## 4. Discussion

In this target trial emulation of 21,636 Canadian middle-aged and older adults, we estimated that a sustained strategy of consuming plant-based and animal-based protein foods at least 1.5 times/day each would have resulted in faster Timed Up&Go and chair-rise times and greater handgrip strength and ALST over 6 years compared with no intervention. Benefits were greater when these protein targets were achieved within an overall healthy dietary pattern. Estimated outcomes were similar whether animal-based protein food frequency was restricted or not while increasing plant-based protein foods. These findings suggest emphasizing both plant- and animal-based protein foods as complementary components of a healthy dietary pattern, rather than competing alternatives, in dietary strategies to support muscle function and strength.

Few prospective studies have documented the effect of the protein food source or the combination of plant-based and animal-based protein foods within a healthy pattern on muscle function, strength or mass among older adults. However, multiple trials assessed the effects of higher protein intake, either from plant or animal sources on muscle outcomes (32, 33), including muscle mass, strength and physical performance, among others. Most trials used protein supplements, meal replacements, gram-per-kilogram protein targets, or structured plant-versus animal-protein diets, rather than changes in the frequency of consumption of protein foods as emulated in the present study. In a meta-analysis of randomized controlled trials that directly compared plant with animal protein (intervention periods, 4 to 104 weeks; mean, 16 weeks), no difference in muscle mass was observed in adults aged ≥60 years (9 trials) or physical performance (5 trials) (32). However, also in adults aged ≥60 years, a greater consumption of animal-based protein yielded a small increase in lower body strength compared with plant-based protein (standardized mean difference, 0.38; 95%CI: 0.05, 0.71; 11 trials) (32). This effect was not observed in adults aged less than 60 years (32). Another meta-analysis in non-frail community-dwelling older adults showed that, compared with control conditions, protein supplementation yielded a small increase in handgrip strength (standardized mean difference, SMD: 0.58; 95% CI: −0.08, 1.24) and gait speed (SMD, 0.41; 95%CI, −0.04, 0.85) although the 95%CI were also compatible with a negligible decrease (33). No effect was observed for chair-rise performance compared with control (33). In this study, we estimated improvements for Timed Up&Go, chair-rise time, and handgrip strength when both plant- and animal-based protein foods were increased alone and within a healthy dietary pattern compared with no intervention. The observation that increased total protein foods, but not the protein food source, may improve components of muscle function or handgrip strength is consistent with prior trials in older adults (33). Divergence from short-term trial literature (e.g., chair rise, ALST) may partly reflect that effects of dietary change on muscle mass and function may take time to become apparent. The 6-year follow-up in the present study suggests that future studies should consider extended intervention periods.

Minimal clinically important differences for walking speed (0.03-0.05 m/s)(34), handgrip strength (5.0 to 6.5 kg)(35), and Timed Up&Go (≥0.8 s, depending on method up to 1.4 s)(36) were developed from short-term clinical or rehabilitation contexts. In the present study, the largest estimates were below these thresholds. For example, the improvement of 0.01 m/s (95%CI: 0.00, 0.02) in walking speed was below the threshold of a meaningful difference of 0.03 to 0.05 (34). Accordingly, these differences should not be interpreted as clinically meaningful for individuals. Measurement error in the SDQ reduces power to detect relationships and may also attenuate estimates (37), although overestimation is also possible (38). The extent to which the small effect sizes are explained mostly by a small causal effect of dietary changes or due to factors such as measurement error remains unclear. Future nutritional observational studies would require a more comprehensive dietary assessment such as repeated 24-h dietary recalls combined with a food-frequency questionnaire (39).

The main strengths of this study include the large sample size as well as the objective and standardized measures of physical function, handgrip strength, and ALST. In addition, the target trial emulation framework and use of the parametric g-formula allowed us to estimate the effects of a sustained dietary intervention strategy while appropriately accounting for time-varying dietary exposures and confounding. Of note, interpreting effect estimates causally hinge on strong assumptions including no unmeasured confounding, no measurement error and no model misspecification. Limitations must also be addressed. First, dietary habits were assessed using a non-quantitative frequency questionnaire, the SDQ. Frequency questionnaires are prone to systematic errors which greatly reduced power to detect differences (40, 41) and can bias estimates (42). Effect estimates should be interpreted cautiously, particularly for chair rise time, walking speed and ALST given their small magnitude. Plus, total dietary intakes could not be assessed directly using the SDQ, although models included a covariate for the total frequency of food consumption as well as body size and physical activity (28). Second, the sensitivity analysis for unmeasured confounding showed that sex-specific estimates may be prone to bias. For example, a varying propensity towards health-seeking behavior was not directly accounted for in the models. This potential confounding phenomenon might contribute to stronger bias in the sex-stratified analysis. Also, the assumption of no unmeasured confounding cannot be confirmed in an observational study. Third, close to half of the sample (41.6%) could not attend the end of follow-up outcome measurement due to COVID-19 protective measures in place at the time. The lower sample size at the end of follow-up has contributed to reducing the precision of effect estimates (wider confidence intervals). However, the limited participation due to COVID-19 protective measures is unlikely to be a major source of bias, since the main reason for having a follow-up during COVID-19 was the date of the baseline assessment. Accordingly, the characteristics of participants who had the opportunity to have their outcome measured are not expected to differ strongly than those who did not have their outcome measured once the COVID-19 pandemic began. Finally, our results may not be generalizable to diverse populations with varying socioeconomic background, considering that the proportion of CLSA participants with a university degree above bachelor (23%) was higher than the national average.

In conclusion, this study aimed to estimate the effect of increasing plant- and animal-based protein foods, alone or within a healthy dietary pattern, as well as the added value of limiting animal-based protein foods among middle-aged and older adults. Results highlight that a combined approach of increasing the frequency of consumption of protein from both plant and animal protein foods yielded modest benefits for muscle function and strength compared with no dietary intervention. The differences were greater when the frequency of consumption of protein foods was increased along with an increased frequency of consumption of vegetables and fruit and whole grain foods. These small effects estimate also hinge on strong assumptions, such as the absence of unmeasured confounding, since they are based on nonexperimental observational data. Thus, experimental studies are required to confirm these findings, as well as studies using dietary data of higher quality.

## Data Availability

Data are available from the Canadian Longitudinal Study on Aging (https://www.clsa-elcv.ca) for researchers who meet the criteria for access to de-identified CLSA data. Analysis code will be made available at https://github.com/didierbrassard/CLSA_TargetTrial_Protein

## Acknowledgements

The opinions expressed in this manuscript are the author’s own and do not reflect the views of the Canadian Longitudinal Study on Aging. The development, testing and validation of the Short Diet Questionnaire (SDQ) were carried out among NuAge study participants as part of the Canadian Longitudinal Study on Aging (CLSA) Phase II validation studies, Canadian Institutes of Health Research (CIHR) 2006-2008. The NuAge study was supported by the Canadian Institutes for Health Research (CIHR), Grant number MOP-62842, and the Quebec Network for Research on Aging, a network funded by the Fonds de Recherche du Québec-Santé.

## Ethics approval

The use of data from the CLSA for this project is approved by the Montreal Heart Institute ethics committee (#2025-3517).

