## Supplemental material for "Estimated effects of plant- and animal-based protein food strategies on muscle function, strength and lean mass after 6 years: a target trial emulation in the Canadian Longitudinal Study on Aging"

### Supplemental Methods

#### Study design: Target trial emulation

**Supplemental Table 1**. Emulation of a dietary intervention target trial using non-experimental data from the CLSA study*^1^*

| **Trial component** | **Target trial specification** | **Target trial emulation** |
| --- | --- | --- |
| **Eligibility criteria** | Participants must live within 50 km of 11 data collection sites, to speak French or English, and to be free of cognitive impairment that would preclude providing informed consent. Participants are not eligible if they have multiple sclerosis, Alzheimer or Parkinson disease, had a surgery in the last 3 months, had polio, had chemotherapy in the last 4 weeks, have a traumatic brain injury with memory problems, have post-traumatic stress disorder or dialysis. | Same.  • Participants were also required to have complete covariate and diet data at baseline.  • Participants were also required data for all **baseline** values of outcome. |
| **Intervention*^2^*** | Each individual is assigned to 1 of the 7 following strategies: 1) habitual diet, i.e., typical Canadian diet; 2) increasing plant-based protein foods; 3) increasing animal-based protein foods; 4) increasing both plant- and animal-based protein foods with balanced targets; 5) increasing only plant-based protein foods, while limiting animal-based protein foods; 6) increasing both plant- and animal-based protein foods, but also vegetables and fruits and whole grain foods; 7) same as 6, but limiting animal-based protein foods. Each strategy is followed until the end of follow-up. Participants assigned to a diet strategy are expected to maintain their dietary habits within the prespecified thresholds for each intervention strategy. | Same.  • We assumed that dietary assessment in the past year also reflected the dietary habits between follow-ups (3-year intervals).  • We assumed that measurement error in dietary intakes was non differential (i.e., did not bias effect estimates). |
| **Assignment** | Participants are randomly assigned to a dietary strategy but are not blinded to their assignment. | Randomization is **not** possible.  • We attempted to emulate randomized assignment by adjusting for baseline variables and baseline value of outcome. The latter aimed to account for confounding effects of cumulative pre-baseline dietary habits and lifestyle on outcome.  • We also assumed that 1) the baseline value of outcome act as a mediator of the effect of dietary habits on outcome; and 2) confounding between the baseline value of outcome and end of follow-up outcome (eg, genetic robustness, “anabolic resistance”) is negligible once the other covariates have been considered. |
| **Outcomes** | Muscle function (Timed Up and Go, time for 1 chair rise 4-m walking speed), strength (handgrip strength), mass (appendicular lean soft tissue mass). In the target trial, the Timed Up and Go would be the primary outcome. | Same. However, there is no primary outcome in the emulation. The aim is not to *detect* an effect, but rather to *estimate* effects for all relevant outcomes. |
| **Time zero and follow-up** | The study starts at baseline and ends at incomplete follow-up or 6 years after baseline, whichever occurs first. Participants are **allowed to discontinue** the intervention when developing incident condition listed in exclusion criteria. | Same.  • An incomplete follow-up was defined as missing data for questionnaires (non-response or loss to follow-up).  • Participants were **excused** from adhering to the intervention when developing incident condition listed in exclusion criteria. |
| **Causal contrast** | • Intention-to-treat effect  • Per-protocol effect | Observational analog of the per-protocol effect. The per protocol effect is the primary contrast of interest. |
| **Statistical analysis** | • Intention-to-treat analysis: apply inverse probability weighting with adjustment for pre- and baseline factors associated with incomplete follow-up to account for incomplete follow-up  • Per-protocol analysis: apply the parametric g-formula algorithm to compare post-intervention outcomes between groups receiving each treatment strategy with adjustment for pre- and postbaseline factors associated with adherence to intervention strategies and incomplete follow-up. | Same. The per protocol effect is the primary contrast of interest.  • The observational analog required additional adjustment for confounding at baseline and before baseline due to prior dietary habits |
| *^1^*CLSA, Canadian Longitudinal Study on Aging. | | |
| *^2^*See 'Hypothetical dietary intervention strategy' in the main text for detailed intervention. | | |

#### Outcomes: Source of data and details of methods of assessment

**Supplemental Table 2.** Outcome and measurement procedures in the Canadian Longitudinal Study on Aging

| **Outcomes** | **Variable name** | **Description and measurement*^1^*** |
| --- | --- | --- |
| Timed Up & Go, seconds | TUG_TIME | Participants were asked to rise from a chair, walk 3 meters, turn around, walk back, and sit down. Participants were allowed to use their assistive device. Participants unable to rise from a chair, stand or walk without the assistance of another person were not eligible. |
| Average time for 1 chair rise, seconds | CR_AVG_TIME | Participants were asked to sit and stand-up five times without using their hands, as quickly as possible. Only valid repetitions were counted. Participants unable to stand or rise from a chair unassisted were not eligible. |
| Walking speed over 4 meters, m/s | WLK_TIME | Participants were asked to walk at their typical pace over a 4-meters course. Participants were allowed to use their assistive device. Participants unable to stand or walk without the assistance of another person were not eligible. |
| Handgrip strength, kg | GS_EXAM_MAX | Handgrip strength was measured using a handheld dynamometer (Tracker Freedom Wireless Grip). Participants were asked to squeeze the dynamometer with as much force as possible. Three contractions were recorded on each hand while sitting. The highest value in all attempts was used. Participants who had a hand surgery in the last 3 months or those who had pain, cast, wounds or prosthetic arms on both sides were not eligible. |
| Appendicular lean soft tissue mass, kg | DXA_WBC_LARM_LEAN,  DXA_WBC_RARM_LEAN,  DXA_WBC_L_LEG_LEAN,  DXA_WBC_R_LEG_LEAN,  DXA_WB_LARM_BMC,  DXA_WB_RARM_BMC,  DXA_WB_LLEG_BMC,  DXA_WB_RLEG_BMC | Calculated as the sum of the upper and lower limbs lean mass minus the sum of the upper and lower limbs bone mineral content, as measured using dual-energy X-ray absorptiometry (DEXA; Hologic Discovery A). Participants with body weight or height exceeding 204 kg and 1.88 m respectively, as well as those who had exposition to an x-ray with contrast (past 2 days) or participated in a nuclear medicine study (past 7 days) were not eligible. |
| *^1^*Participants were allowed one practice trial before conducting the tests. | | |

#### Statistical analyses: Modelling approach for each variable

*Supplemental* Table 3 shows how each variable was modeled in the g-formula algorithm.

**Supplemental Table 3**. Modeling approach for each variable in the g-formula algorithm

| **Variable** | **Modeling as independent variable** | **Time-varying variable** | **Modeling as dependent variable** |
| --- | --- | --- | --- |
| **Covariates** | | | |
| Age | Continuous: restricted cubic spline with 5 knots | No | Not applicable |
| Sex | Dichotomous | No | Not applicable |
| Height | Continuous: restricted cubic spline with 4 knots | No | Not applicable |
| Baseline value of outcome | Continuous: restricted cubic spline with 5 knots | No | Not applicable |
| Education level | Categorical | No | Not applicable |
| Province of residence | Categorical | No | Not applicable |
| Supplement use (vitamin, mineral, omega 3-6-9, or natural health product) | Dichotomous | No | Not applicable |
| Former smoking | Dichotomous | No | Not applicable |
| Fat sources | Continuous: linear | Yes | Normal (linear regression) |
| Total foods | Continuous: restricted cubic spline with 4 knots | Yes | Normal (linear regression) |
| Bodyweight | Continuous: restricted cubic spline with 5 knots | Yes | Normal (linear regression) |
| Menopausal status (female only) | Dichotomous | Yes | Binomial (logistic regression) |
| Hormone replacement therapy use (female only) | Dichotomous | Yes | Binomial (logistic regression) |
| Marital status | Categorical | Yes | Binomial (logistic regression) |
| Social participation | Continuous: restricted cubic spline with 4 knots | Yes | Normal (linear regression) |
| Living alone | Dichotomous | Yes | Binomial (logistic regression) |
| Retirement status | Dichotomous | Yes | Binomial (logistic regression) |
| High blood pressure | Dichotomous | Yes | Binomial (logistic regression) |
| Diabetes | Dichotomous | Yes | Binomial (logistic regression) |
| Cancer diagnosis | Dichotomous | Yes | Binomial (logistic regression) |
| Current smoker | Dichotomous | Yes | Binomial (logistic regression) |
| Frequency of alcohol consumption in the past 12 months | Categorical | Yes | Binomial (logistic regression) |
| Sitting time in the past week | Continuous: restricted cubic spline with 4 knots | Yes | Normal (linear regression) |
| Light intensity or greater physical activity in the past week | Continuous: restricted cubic spline with 4 knots | Yes | Zero-inflated normal (linear regression) |
| Compliance excused at follow-ups | Dichotomous | Yes | Binomial (logistic regression) |
| **Intervention variables** | | | |
| Protein foods, plant-based (incl. beverages) | Continuous: restricted cubic spline with 4 knots | Yes | Normal (linear regression) |
| Protein foods, animal-based (incl. milk) | Continuous: restricted cubic spline with 4 knots | Yes | Normal (linear regression) |
| Vegetables and fruits | Continuous: restricted cubic spline with 5 knots | Yes | Normal (linear regression) |
| Whole grains | Continuous: restricted cubic spline with 3 knots | Yes | Normal (linear regression) |

### Supplemental Results

#### Characteristics of non eligible participants

**Supplemental Table 4**. Characteristics of CLSA participants at baseline, by eligibility status*^1^*

| **Characteristic** | **Eligible** N = 21,636*^2^* | **Non-eligible** N = 8,461*^2^* |
| --- | --- | --- |
| Female sex | 10,571 (49%) | 4,749 (56%) |
| Age, y | 62 (54, 70) | 64 (56, 73) |
| Education level |  |  |
| Elementary | 2,963 (14%) | 1,536 (18%) |
| High school or trade school | 3,769 (17%) | 1,812 (21%) |
| College | 3,700 (17%) | 1,465 (17%) |
| University certificate or bachelor's degree | 6,257 (29%) | 2,084 (25%) |
| University degree above bachelor | 4,947 (23%) | 1,557 (18%) |
| Missing | 0 | 7 |
| Body mass index, kg/m² | 27.1 (24.3, 30.5) | 27.6 (24.6, 31.4) |
| Missing | 0 | 136 |
| Body mass index category |  |  |
| <18.5 kg/m² | 143 (0.7%) | 74 (0.9%) |
| 18.5 to <25 kg/m² | 6,582 (30%) | 2,282 (27%) |
| 25 to <30 kg/m² | 8,878 (41%) | 3,210 (39%) |
| 30 kg/m² or more | 6,033 (28%) | 2,759 (33%) |
| Missing | 0 | 136 |
| Menopausal status | 8,593 (81%) | 4,067 (86%) |
| Missing | 11,065 | 3,736 |
| Ever taken hormone replacement therapy | 3,839 (36%) | 1,909 (40%) |
| Missing | 11,065 | 3,737 |
| Marital status |  |  |
| Single | 1,798 (8.3%) | 856 (10%) |
| Previously partnered | 4,395 (20%) | 2,389 (28%) |
| Married or common-law | 15,443 (71%) | 5,208 (62%) |
| Missing | 0 | 8 |
| Frequency of social participation activities per day | 0.39 (0.22, 0.64) | 0.36 (0.21, 0.64) |
| Living alone | 4,485 (21%) | 2,337 (28%) |
| Missing | 0 | 27 |
| Completely or partly retired | 11,578 (54%) | 5,104 (60%) |
| Missing | 0 | 5 |
| Has high blood pressure | 7,584 (35%) | 3,517 (42%) |
| Missing | 0 | 64 |
| Has diabetes | 3,452 (16%) | 1,858 (22%) |
| Missing | 0 | 61 |
| Has had cancer | 3,075 (14%) | 1,562 (19%) |
| Missing | 0 | 61 |
| Taking dietary supplement or natural health product | 11,855 (55%) | 4,959 (59%) |
| Smoking status |  |  |
| Never | 6,829 (32%) | 2,671 (32%) |
| Former | 13,130 (61%) | 4,866 (58%) |
| Current | 1,677 (7.8%) | 899 (11%) |
| Missing | 0 | 25 |
| Frequency alcohol consumption past 12 months |  |  |
| Never | 2,269 (10%) | 1,168 (15%) |
| <1 time a month to 1 time a month | 3,854 (18%) | 1,826 (24%) |
| >1 time a month to 1 time a week | 4,715 (22%) | 1,576 (20%) |
| >1 time a week | 10,798 (50%) | 3,175 (41%) |
| Missing | 0 | 716 |
| Sitting time, minutes per day, last 7 days | 154 (154, 206) | 154 (154, 206) |
| Missing | 0 | 1,311 |
| Physical activity of light intensity of higher, minutes per day, last 7 days | 16 (0, 46) | 10 (0, 39) |
| Missing | 0 | 1,313 |
| *^1^*CLSA, Canadian Longitudinal Study on Aging | | |
| *^2^*n (%); Median (Q1, Q3) | | |

#### Observed (crude values) and missing diet and time-varying covariate data during follow-up

**Supplemental Table 5.** Observed values of dietary assessment and time-varying covariates across follow-up among 21,636 eligible CLSA participants*^1^*

| **Characteristic** | **Baseline** N = 21,636*^2^* | **Follow-up 1** N = 21,636*^2^* | **End of follow-up** N = 21,636*^2^* |
| --- | --- | --- | --- |
| Plant-based protein foods incl. beverages, freq/d | 0.86 (0.43, 1.29) | 0.86 (0.43, 1.29) | 0.86 (0.43, 1.29) |
| Missing | 0 | 886 | 2,357 |
| Animal-based protein foods incl. milk, freq/d | 2.96 (2.24, 3.71) | 2.75 (2.10, 3.46) | 2.71 (2.02, 3.39) |
| Missing | 0 | 886 | 2,357 |
| Vegetables and fruits, freq/d | 3.43 (2.43, 4.57) | 3.14 (2.32, 4.14) | 3.14 (2.29, 4.14) |
| Missing | 0 | 886 | 2,357 |
| Whole grain foods, freq/d | 1.07 (0.71, 1.57) | 1.00 (0.57, 1.43) | 1.00 (0.46, 1.43) |
| Missing | 0 | 886 | 2,357 |
| Added fat, freq/d | 1.03 (0.57, 1.43) | 1.07 (0.57, 1.43) | 1.07 (0.57, 1.43) |
| Missing | 0 | 886 | 2,357 |
| Total foods and beverages, freq/d | 11.4 (9.4, 13.6) | 10.8 (9.0, 13.0) | 11.0 (9.0, 13.1) |
| Missing | 0 | 886 | 2,357 |
| Body weight, kg | 78 (67, 89) | 78 (67, 89) | 77 (67, 89) |
| Missing | 0 | 1,294 | 2,740 |
| Menopausal status (female only) | 8,593 (40%) | 9,049 (44%) | 8,942 (47%) |
| Missing | 0 | 1,019 | 2,493 |
| Hormone replacemement therapy use (female only) | 3,839 (18%) | 4,132 (20%) | 4,129 (22%) |
| Missing | 0 | 933 | 2,444 |
| Social participation, freq/d | 0.39 (0.22, 0.64) | 0.36 (0.21, 0.61) | 0.33 (0.18, 0.57) |
| Missing | 0 | 886 | 2,357 |
| Living alone | 4,485 (21%) | 4,554 (22%) | 4,457 (24%) |
| Missing | 0 | 1,242 | 2,690 |
| Completely or partly retired from work | 11,578 (54%) | 12,741 (62%) | 13,242 (69%) |
| Missing | 0 | 1,070 | 2,461 |
| Has high blood pressure | 7,584 (35%) | 7,490 (37%) | 7,448 (40%) |
| Missing | 0 | 1,557 | 2,833 |
| Has diabetes | 3,452 (16%) | 3,691 (18%) | 3,719 (20%) |
| Missing | 0 | 1,557 | 2,834 |
| Had cancer history or has cancer | 3,075 (14%) | 3,531 (18%) | 3,519 (19%) |
| Missing | 0 | 1,557 | 2,750 |
| Current smoker | 1,677 (7.8%) | 1,362 (6.6%) | 1,025 (5.3%) |
| Missing | 0 | 889 | 2,414 |
| Sitting time in the past week, min/day | 154 (154, 206) | 206 (154, 206) | 206 (154, 206) |
| Missing | 0 | 890 | 2,360 |
| Light intensity or greater physical activity in the past week, min/day | 16 (0, 46) | 19 (0, 48) | 13 (0, 42) |
| Missing | 0 | 890 | 2,360 |
| Marital status |  |  |  |
| Single | 1,798 (8.3%) | 1,783 (8.6%) | 1,545 (8.0%) |
| Previously partnered | 4,395 (20%) | 4,385 (21%) | 4,335 (23%) |
| Married or common-law | 15,443 (71%) | 14,572 (70%) | 13,340 (69%) |
| Missing | 0 | 896 | 2,416 |
| *^1^*CLSA, Canadian Longitudinal Study on Aging | | | |
| *^2^*Median (Q1, Q3); n (%) | | | |

#### Proportion of participants meeting dietary intervention targets

**Supplemental Table 6**. Proportion of eligible CLSA participants (21,636) meeting dietary intervention variable thresholds across follow-ups, by hypothetical intervention*^1^*

| **Intervention** | **Number of variables intervened on** | **Baseline** | **Follow-up 1** | **End of follow-up** |
| --- | --- | --- | --- | --- |
| Higher plant protein foods | 1 | 2,404 (11.1%) | 2,050 (9.9%) | 1,813 (9.4%) |
| Higher animal protein foods | 1 | 7,279 (33.6%) | 6,477 (31.2%) | 5,679 (29.5%) |
| Higher total protein foods | 2 | 1,973 (9.1%) | 1,641 (7.9%) | 1,372 (7.1%) |
| Higher plant but limited animal protein foods | 2 | 431 (2.0%) | 409 (2.0%) | 441 (2.3%) |
| Higher total protein foods within a healthy dietary pattern | 4 | 845 (3.9%) | 601 (2.9%) | 498 (2.6%) |
| Higher plant but limited animal protein foods within a healthy dietary pattern | 4 | 160 (0.7%) | 122 (0.6%) | 159 (0.8%) |
| *^1^*Values are n (%). See Table 1 in the main manuscript for detailed intervention targets. | | | | |

#### Observed (crude values) and missing outcome data during follow-up

**Supplemental Table 7**. Observed mean values of muscle function, handgrip strength and appendicular lean soft tissue across follow-up among 21,636 eligible CLSA participants*^1^*

| **Characteristic** | **Baseline** N = 21,636*^2^* | **Follow-up 1** N = 18,279*^2^* | **End of follow-up** N = 17,427*^2^* |
| --- | --- | --- | --- |
| Timed Get 'Up and Go', s | 9.31 (2.01) | 9.67 (2.12) | 10.08 (2.37) |
| Missing | 0 | 552 | 7,413 |
| Walking speed over 4 meters, m/s | 1.00 (0.19) | 0.98 (0.17) | 0.97 (0.18) |
| Missing | 0 | 525 | 7,391 |
| Chair rise, average time for 1 repetition, s | 2.65 (0.76) | 2.54 (0.77) | 2.56 (0.71) |
| Missing | 0 | 927 | 7,696 |
| Maximal handgrip strength across all trials, kg | 36 (12) | 35 (12) | 34 (11) |
| Missing | 0 | 849 | 7,610 |
| Appendicular lean soft tissue, kg | 21.3 (5.6) | 21.6 (5.6) | 21.4 (5.6) |
| Missing | 0 | 1,465 | 7,990 |
| *^1^*CLSA, Canadian Longitudinal Study on Aging | | | |
| *^2^*Mean (SD) | | | |

#### Subgroup analysis, by sex

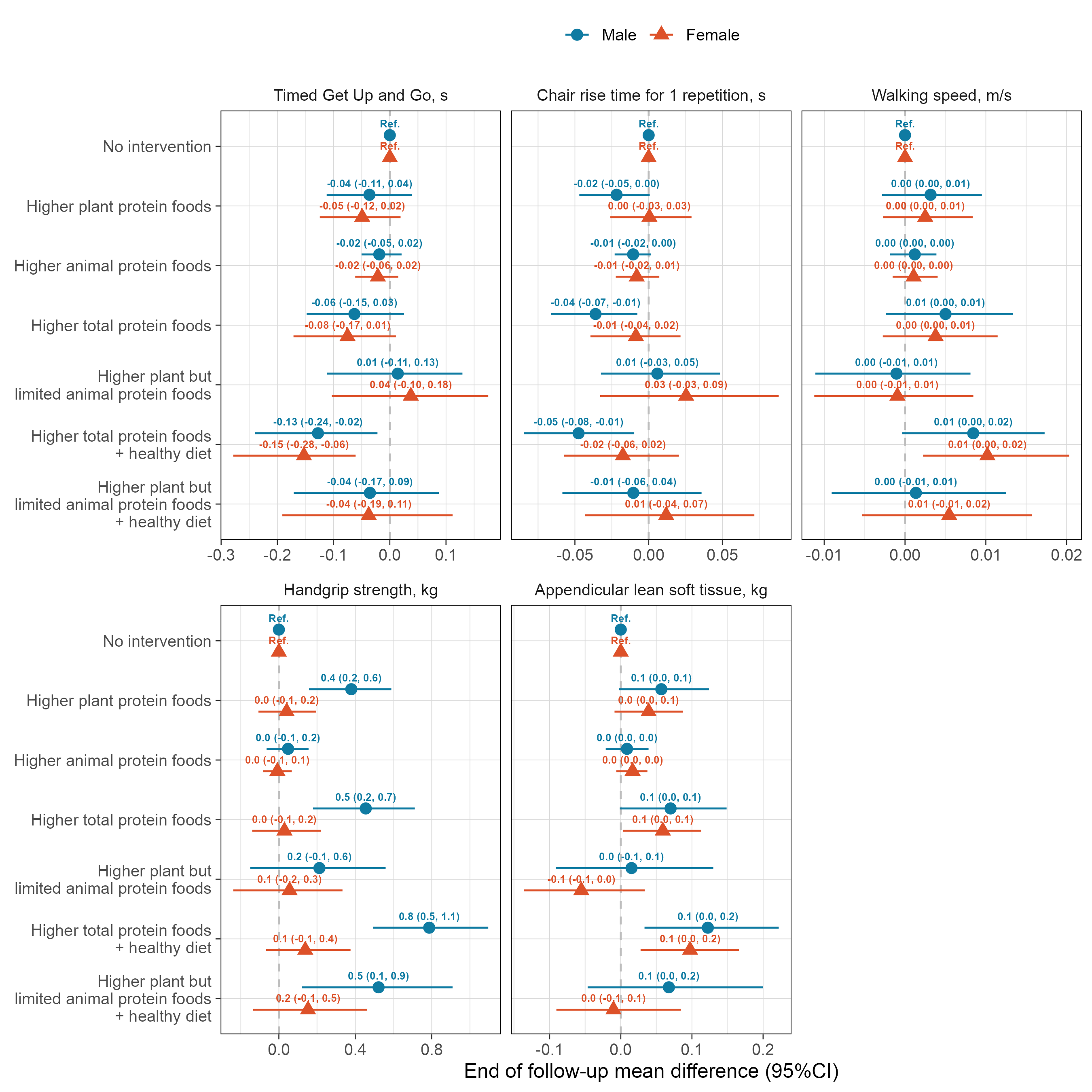

**Supplemental Figure 1**. End-of-follow-up mean differences (95%CI) in muscle function, handgrip strength and appendicular lean soft tissue mass under each 6-year hypothetical dietary interventions, compared with no intervention among eligible CLSA participants (n=21,636), by sex. Effect estimates were obtained using the parametric g-formula algorithm. Models were adjusted for the covariates age, height, baseline value of outcome, education level, province of residence, supplement use (vitamin, mineral, omega 3-6-9, or natural health product), former smoking, fat sources, total foods, bodyweight, menopausal status (female only), hormone replacement therapy use (female only), marital status, social participation, living alone, retirement status, high blood pressure, diabetes, cancer diagnosis, current smoker, frequency of alcohol consumption in the past 12 months, sitting time in the past week, as well as light intensity or greater physical activity in the past week. CLSA, Canadian Longitudinal Study on Aging

#### Subgroup analysis, by age category

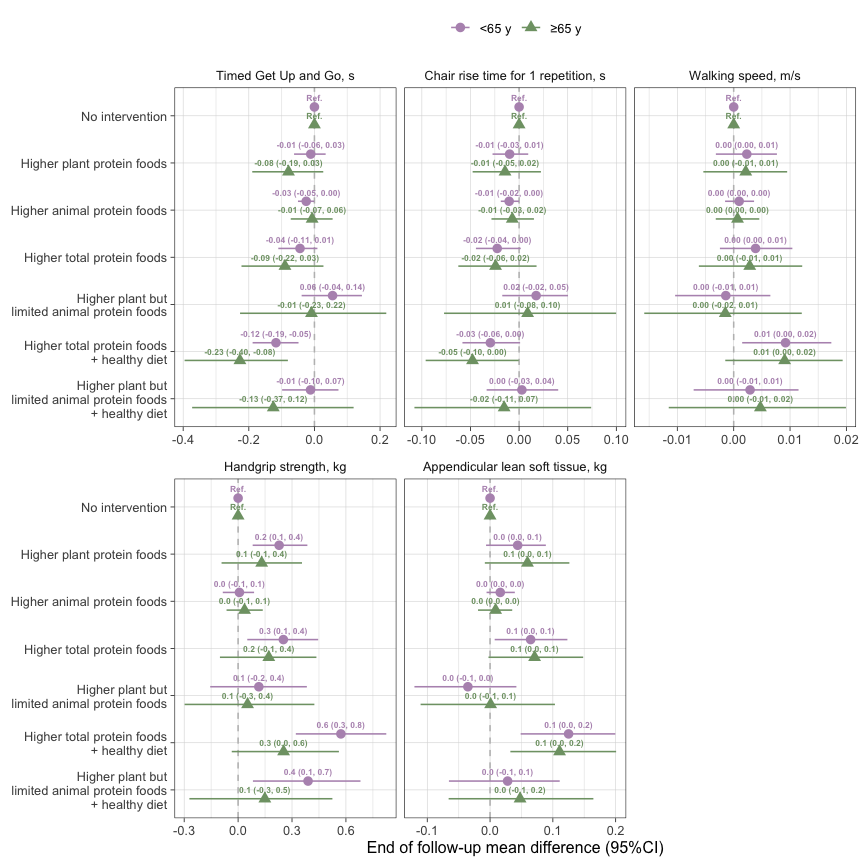

***Supplemental Figure 2:*** *Mean difference in muscle function, grip strength and appendicular lean soft tissue after the 6-year hypothetical interventions, compared with no intervention, in eligible CLSA participants, by age group. Effect estimates were obtained using the g-formula algorithm (see Methods). Models were adjusted for the covariates sex, height, baseline value of outcome, education level, province of residence, supplement use (vitamin, mineral, omega 3-6-9, or natural health product), former smoking, fat sources, total foods, bodyweight, menopausal status (female only), hormone replacement therapy use (female only), marital status, social participation, living alone, retirement status, high blood pressure, diabetes, cancer diagnosis, current smoker, frequency of alcohol consumption in the past 12 months, sitting time in the past week, as well as light intensity or greater physical activity in the past week. CFG, Canada’s Food Guide; CLSA, Canadian Longitudinal Study on Aging.*

#### Sensitivity Analyses

##### Assessment of unmeasured confounding

*Supplemental* Table 8 presents the estimated effects of the hypothetical intervention on end of follow-up probability of injury serious enough to limit daily activities in the past 12 months. Accidental fall from the same level were excluded to avoid including fall that could be prevented by maintaining proper nutritional status. However, accidental fall from height were included. Accidental injury should not be strongly influenced by dietary intakes.

**Supplemental Table 8**. Estimated effects of the hypothetical intervention on end of follow-up probability of serious injury in the past 12 months among eligible participants (n=21,636)*^1^*

| Hypothetical intervention | Probability of injury | Probability ratio (95%CI) |
| --- | --- | --- |
| **All (n=21636)** | | |
| No intervention | 12.4% | Ref. |
| Higher plant protein foods | 12.6% | 1.01 (0.96, 1.07) |
| Higher animal protein foods | 12.4% | 1.00 (0.97, 1.03) |
| Higher total protein foods | 12.6% | 1.02 (0.94, 1.09) |
| Higher plant but limited animal protein foods | 12.9% | 1.04 (0.94, 1.13) |
| Higher total protein foods within a healthy dietary pattern | 11.8% | 0.95 (0.88, 1.03) |
| Higher plant but limited animal protein foods within a healthy dietary pattern | 12.2% | 0.98 (0.88, 1.07) |
| **Males (n=11065)** | | |
| No intervention | 13.6% | Ref. |
| Higher plant protein foods | 13.4% | 0.98 (0.91, 1.06) |
| Higher animal protein foods | 13.0% | 0.96 (0.91, 1.00) |
| Higher total protein foods | 12.7% | 0.94 (0.85, 1.04) |
| Higher plant but limited animal protein foods | 15.1% | 1.11 (0.99, 1.24) |
| Higher total protein foods within a healthy dietary pattern | 12.2% | 0.90 (0.80, 1.01) |
| Higher plant but limited animal protein foods within a healthy dietary pattern | 14.2% | 1.04 (0.90, 1.19) |
| **Females (n=10571)** | | |
| No intervention | 11.4% | Ref. |
| Higher plant protein foods | 12.0% | 1.05 (0.96, 1.12) |
| Higher animal protein foods | 11.9% | 1.04 (1.01, 1.08) |
| Higher total protein foods | 12.6% | 1.10 (0.99, 1.21) |
| Higher plant but limited animal protein foods | 10.9% | 0.95 (0.83, 1.08) |
| Higher total protein foods within a healthy dietary pattern | 12.0% | 1.06 (0.93, 1.17) |
| Higher plant but limited animal protein foods within a healthy dietary pattern | 10.7% | 0.94 (0.80, 1.07) |
| **Age<65 y (n=12991)** | | |
| No intervention | 15.1% | Ref. |
| Higher plant protein foods | 15.3% | 1.02 (0.95, 1.09) |
| Higher animal protein foods | 14.9% | 0.99 (0.96, 1.02) |
| Higher total protein foods | 15.2% | 1.01 (0.92, 1.10) |
| Higher plant but limited animal protein foods | 16.3% | 1.08 (0.97, 1.19) |
| Higher total protein foods within a healthy dietary pattern | 14.3% | 0.95 (0.86, 1.05) |
| Higher plant but limited animal protein foods within a healthy dietary pattern | 15.4% | 1.02 (0.91, 1.15) |
| **Age≥65 y (n=8645)** | | |
| No intervention | 8.3% | Ref. |
| Higher plant protein foods | 8.4% | 1.01 (0.90, 1.14) |
| Higher animal protein foods | 8.6% | 1.03 (0.96, 1.10) |
| Higher total protein foods | 8.7% | 1.04 (0.90, 1.20) |
| Higher plant but limited animal protein foods | 7.6% | 0.91 (0.68, 1.12) |
| Higher total protein foods within a healthy dietary pattern | 7.9% | 0.94 (0.77, 1.09) |
| Higher plant but limited animal protein foods within a healthy dietary pattern | 7.1% | 0.86 (0.62, 1.06) |
| *^1^*Values were estimated using the parametric g-formula (see Methods). 'No intervention' corresponds to the estimated probability of injury when dietary intakes are not modified. Variance was estimated using 500 bootstrap resamples. Sex- and age-specific estimates were obtained by stratification. Models were adjusted for age, sex (except for sex-stratified models), height, baseline value of outcome, education level, province of residence, supplement use (vitamin, mineral, omega 3-6-9, or natural health product), former smoking, fat sources, total foods, bodyweight, menopausal status (female only), hormone replacement therapy use (female only), marital status, social participation, living alone, retirement status, high blood pressure, diabetes, cancer diagnosis, current smoker, frequency of alcohol consumption in the past 12 months, sitting time in the past week, as well as light intensity or greater physical activity in the past week. Age (continuous) was kept in the age-stratified models to account for residual confounding within age stratum. CFG, Canada's Food Guide. | | |

##### Assessment of model misspecification

*Supplemental* Figure 3 presents the comparison between observed (“IP weighted estimates”; natural course accounting for loss to follow-up) and predicted values (“parametric g formula estimates”) of time-varying variables.

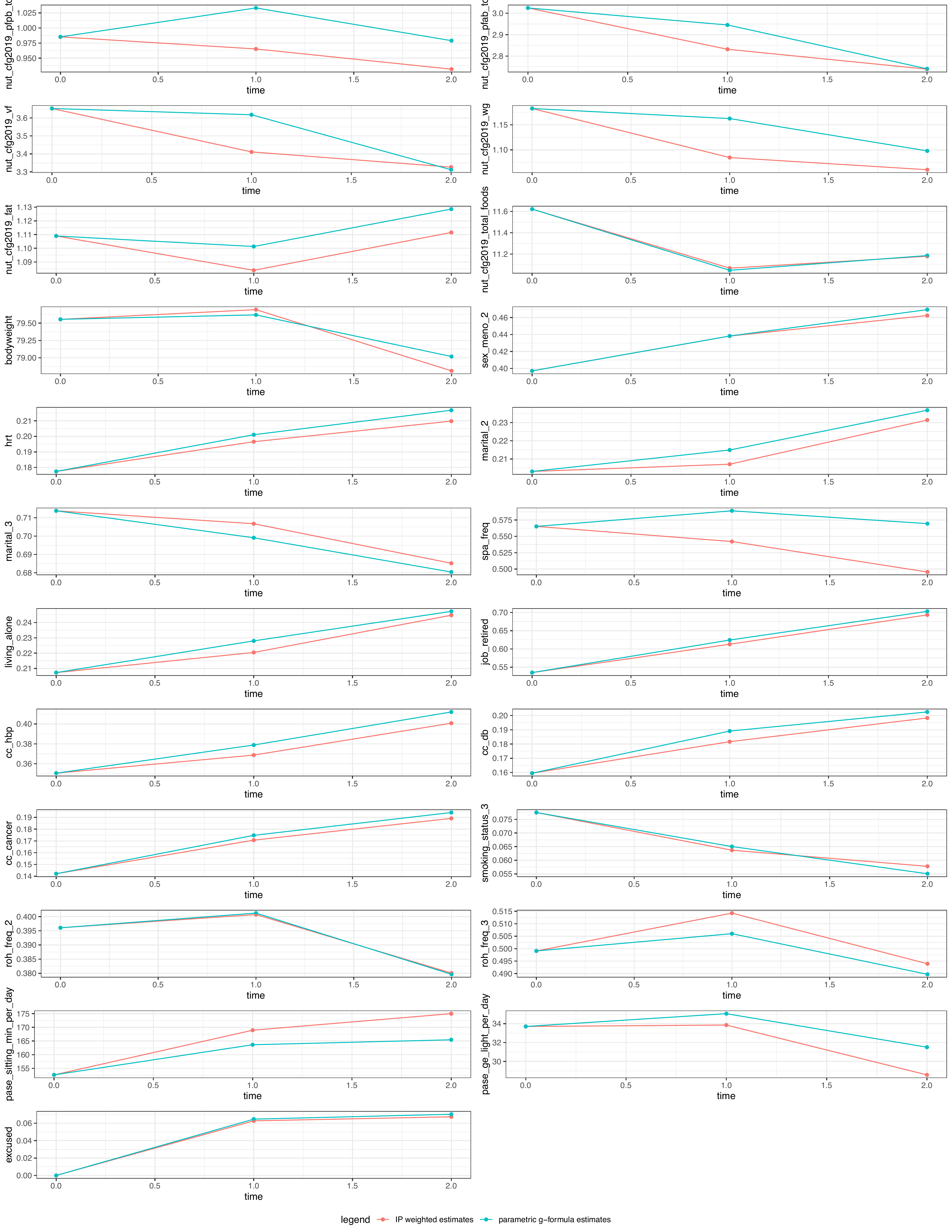

**Supplemental Figure 3.** Assessment of time-varying models misspecification in the g formula algorithm. Divergence between the parametric g−formula estimates and the inverse-probability weighted estimates suggest that the functional form of covariates and/or modelling of outcomes may have been incorrect. Variables definition: nut_cfg2019_pfpb, protein foods, plant-based; nut_cfg2019_pfab, protein foods, animal-based; nut_cfg2019_vf, vegetables and fruits; nut_cfg2019_wg, whole grains; nut_cfg2019_fat, fats; nut_cfg2019_total_foods, total foods; bodyweight, bodyweight; sex_meno_2, menopaused; hrt, hormone replacement therapy use; marital_2, marital status; marital_3, marital status; spa_freq, social participation; living_alone, living alone; job_retired, completely or partly retired; cc_hbp, has high blood pressure; cc_db, has diabetes; cc_cancer, had a cancer diagnosis; smoking_status_3, smoking status; roh_freq_2, frequency of alcohol consumption in the past 12 months; roh_freq_3, frequency of alcohol consumption in the past 12 months; pase_sitting_min_per_day, sitting time in the past week; pase_ge_light_per_day, light intensity or greater physical activity in the past week; excused, compliance excused at follow-ups.
